# Hip and knee joint kinematics predict quadriceps hyperreflexia in people with post-stroke Stiff-Knee gait

**DOI:** 10.1101/2022.11.09.22282134

**Authors:** Jeonghwan Lee, Tunc Akbas, James Sulzer

## Abstract

Wearable assistive technology for the lower extremities has shown great promise towards improving gait function in people with neuromuscular injuries. But common secondary impairments, such as hyperreflexia, have been often neglected. Adding hyperreflexia prediction to the control loop would require expensive or complex measurement of muscle fiber characteristics. In this study, we explore a clinically accessible biomechanical predictor set that can accurately predict rectus femoris (RF) reaction after knee flexion assistance in pre-swing by a powered orthosis. We examined a total of 14 gait parameters based on gait kinematic, kinetic, and simulated muscle-tendon states from 8 post-stroke individuals with Stiff-Knee gait (SKG) wearing a knee exoskeleton robot. We independently performed both parametric and non-parametric variable selection approaches using machine learning regression techniques. Both models revealed the same four kinematic variables relevant to knee and hip joint motions were sufficient to effectively predict RF hyperreflexia. These results suggest that control of knee and hip kinematics may be a more practical method of incorporating quadriceps hyperreflexia into the exoskeleton control loop than the more complex acquisition of muscle fiber properties.

## Introduction

Gait disorders are common in stroke survivors. More than 80% of stroke survivors have varying degrees of gait abnormalities^11,21^, and about 25% have a residual impairment that requires full physical assistance, despite rehabilitation efforts^17^. Recent technological advances have introduced wearable robots as potential solutions to mitigate physical barriers for people with impaired mobility. There are various commercialized and research purpose lower-body exoskeletons showing promise in performance augmentation, mobility assistance, and gait therapy following neurological injuries^12,41^. Assistive strategies using exoskeletons incorporate force, torque, electromyography (EMG), and gait kinematics into the control loop^22,41^. More recently, human-in-the-loop approaches have been introduced to maximize the benefit of the assistive device with feedback of metabolic energy measures in a real-time optimization^43^. However, existing exoskeletons have only addressed weakness and ignored critical neuromuscular impairments, such as hyperreflexia, defined as a hypersensitive stretch reflex, governed by musculotendon fiber stretch velocity^42^.

Stiff-Knee gait (SKG) is a common abnormal gait pattern following stroke, characterized by diminished knee flexion during the swing phase of the gait cycle. We previously developed a lightweight, remotely actuated powered knee orthosis^38^ to assist post-stroke individuals with SKG. This robotic knee exoskeleton was expected to improve swing-phase knee flexion kinematics. While kinematics improved somewhat, hyperreflexia in the rectus femoris (RF), represented by excessive early swing-phase muscle activation, primarily followed exoskeletal assistance, but occasionally without assistance as well^2^. Musculoskeletal modeling and simulation revealed that increased RF fiber stretch velocity preceded increases in RF muscle activation^2^. In a new cohort of individuals with SKG, Akbas et al. found that RF reflex excitability was highly associated with reduced swing phase knee flexion angle^1^. Taken together, this evidence indicates that robotic knee perturbations on post-stroke individuals with SKG could elicit counterproductive quadriceps hyperreflexia, a reaction likely spurred on by suprathreshold RF fiber stretch velocity. Thus, if excessive RF fiber stretch velocity could be avoided, it may be possible to enable the full benefits of exoskeletal assistance in those with post-stroke SKG. However, real-time measurement of fiber stretch velocity requires difficult to access equipment such as ultrasound and a real-time analysis and control system^29^. Another approach could be to identify clinically accessible correlates of fiber stretch velocity to avoid the hyperreflexia elicited by exoskeletal assistance.

The objective of this study was to determine to what degree kinematic and kinetic features could substitute for RF fiber stretch velocity in predicting hyperreflexive RF muscle activation following exoskeletal assistance. To achieve this, we performed parametric and nonparametric multivariate regression analyses for variable selection on a knee exoskeleton gait data from 8 post-stroke participants with SKG^38^. We hypothesized that hip and knee flexion kinematics would predict RF hyperreflexia due to the biarticular characteristics of the RF. This work represents a novel statistical and machine learning regression approach to identify predictors of hyperreflexia. Obtaining clinically accessible predictors for hyperreflexia will lead to more effective human-in-the-loop assistive strategies that account for neural impairments.

## Materials and Methods

### Experimental Data

We obtained previously collected data from 8 chronic, hemiparetic participants with post-stroke SKG who gave written informed consent using procedures approved by the local Institutional Review Board^37^. Inclusion criteria for the hemiparetic participants included their peak paretic knee flexion during swing was at least 15° less than their peak nonparetic knee flexion and the ability to walk for 20 min without rest at 0.55 m/s on a treadmill. A lightweight, powered knee orthosis was used to provide knee flexion torque perturbations during the pre-swing phase without affecting the remainder of the gait cycle^38^. Analog data, including GRF from the instrumented split-belt treadmill (Tecmachine, Andrez Boutheon, France) and applied torque by the powered orthosis, were acquired at 1 kHz. Motion capture data (Motion Analysis, Santa Rosa, CA) on the lower limbs was collected at 120 Hz. Electromyography (EMG) (Delsys Inc., Boston, MA) from the RF muscle was measured at 1 kHz.

The experimental protocol involved steps with and without pre-swing knee flexion assistance. Steps with knee flexion assistance ranged from 10 Nm to 40 Nm. A mean of less than 1 Nm of resistance was measured on the knee for the remainder of the gait cycle. The weight of the device did not alter walking kinematics. In this study, a total of 406 gait cycles were extracted including 260 gait cycles with knee flexion assistance (Mean ± SD: 23.32 ± 5.85 Nm; Min: 11.40 Nm; Max 34.51 Nm). Previous work describes further details of the protocol^37^.

### Musculoskeletal Modeling and Simulation

In order to determine muscle-tendon states (i.e., RF muscle fiber stretch velocity at each time instance), we employed musculoskeletal modeling and simulation through OpenSim 4.3^10^ using a method validated in previous work^2,4^. To summarize, we condensed upper body segments in the gait 2392 model of OpenSim to the pelvis segment to account for a lower body marker set, so that it had 18 degrees of freedom and 90 muscle-tendon actuators. The modified model was scaled to match the anthropometry of each subject. Marker data were fed into the inverse kinematics tool, which generated joint kinematics with the least square fit of marker trajectories. A residual reduction algorithm (RRA) adjusted the model to minimize dynamic inconsistencies between the experimental GRFs and body segment kinematics. Based on the adjusted model after RRA, computed muscle control (CMC)^39^ estimated muscletendon states and excitation patterns that reproduced the motion while minimizing the sum of excitations squared. Unmeasured handrail forces were estimated as previously described^3^. The dynamic consistency of the simulations was evaluated by examining the resulting residual forces and moments and verifying them with the OpenSim guidelines^18^.

### Dependent Variable

The dependent variable for the regression analysis was RF activity. The raw RF EMG signals were filtered with a fourth-order band-pass Butterworth filter with cutoff frequencies of 20–400 Hz to remove artifacts, demeaned, rectified, and low-pass filtered with a 4th order Butterworth filter at 10 Hz. To reduce inter-individual variability, we used the mean of the EMG envelope in the trial as the normalization reference^8^. The processed signals were divided into gait cycles based on a paretic limb heel-strike event using vertical GRFs and then normalized into 100-time frames. A reflex response would occur within 120ms after stimulus onset through mono- or polysynaptic mechanisms^31^. This time interval has been employed to detect quadriceps stretch reflexes following mechanical knee perturbations during gait^27^. In addition to the 120ms window, we accounted for timing errors from computation of RF fiber stretch velocity and evaluated windows of 90ms and 150ms. We defined the initiation of reflex as the timing of the peak pre-swing muscle fiber stretch velocity estimated from the musculoskeletal simulation. We then numerically integrated the RF EMG envelopes following stimulus onset to obtain integrated EMG (iEMG) measures representing an involuntary response. **Figure 1** visualizes RF iEMG for involuntary responses.

**Figure 1:**
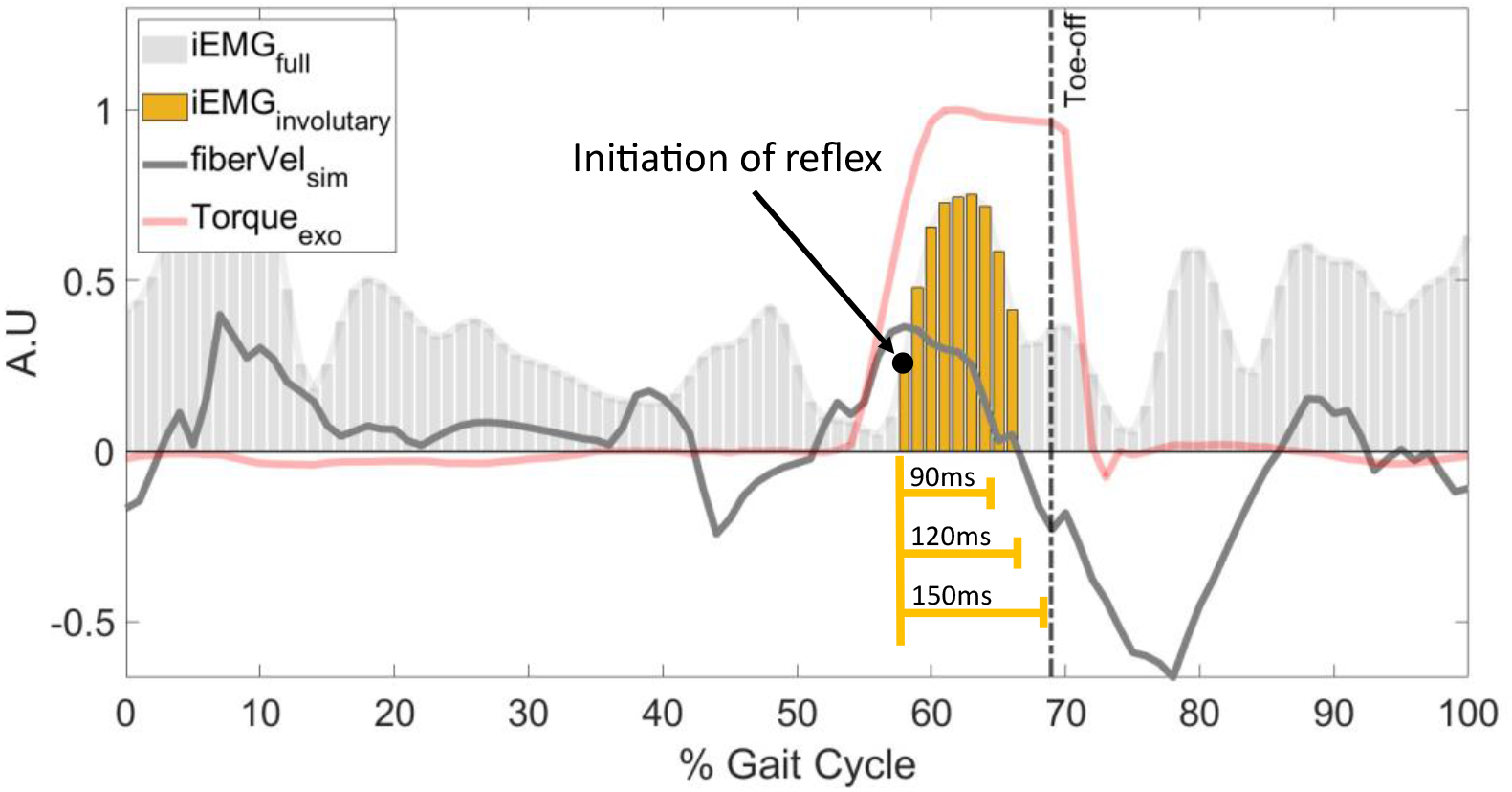
Visualization of the rectus femoris EMG signals, knee flexion torque assistance, and simulated RF muscle fiber stretch velocity. Gray line indicates simulated RF fiber velocity. Red solid line represents the external knee torque assistance profile by the powered knee exoskeleton device. Vertical bars show iEMG of RF. The involuntary response is captured from the peak simulated RF fiber velocity until 120 ± 30ms following (yellow).

### Independent Variables

A total of 14 predictors were extracted from each gait cycle based on muscle-tendon states as well as gait kinematic and kinetic data. One muscle-tendon relevant variable was selected from the musculoskeletal simulation: the peak pre-swing RF fiber stretch velocity. Three variables from hip and knee joint kinematics in the pre-swing phase (i.e., peak hip flexion and knee flexion velocity, and relative peak knee flexion velocity to peak hip flexion) were chosen due to the biarticular physiology of the RF and its associations with post-stroke SKG^15^. Correspondingly, we used body kinematics which are velocities and accelerations based on the configuration (center of mass position and orientation) of each body segment. Specifically, we used 3 body kinematic variables from the thigh and shank segments (i.e., peak thigh and shank angular velocity, and relative peak shank angular velocity to peak thigh) in the pre-swing phase. Additionally, we used the pelvic segment acceleration in the anteroposterior plane as a proxy for body acceleration^7^. The remaining six variables were acquired from propulsive/brake impulse and ground reaction forces. Specifically, we computed the paretic limb’s propulsive, braking, and net impulse during the pre-swing phase ^6^. GRF signals were processed to obtain the peak anterior/posterior GRF in the pre-swing phase, peak medial/lateral GRF in the stance phase, and peak vertical GRF in the stance phase (**Table 1**).

**Table 1:** List of parameters.

| **Description** | **Abbreviation** | **Category** |
| --- | --- | --- |
| Peak rectus femoris fiber stretch velocity in pre-swing<sup>2</sup> (m/s) | RectusFemoris_Vel | Muscle-tendon mechanics |
| Peak hip flexion velocity in pre-swing (rad/s) | HipFlex_Vel | Joint kinematics |
| Peak knee flexion velocity in pre-swing<sup>30</sup> (rad/s) | KneeFlex_Vel | Joint kinematics |
| Relative peak knee flexion velocity with respect to peak hip flexion (rad/s) | relKneeHip_Vel | Joint kinematics |
| Peak pelvic segment anterior/posterior acceleration in pre-swing<sup>7</sup> (m/s<sup>2</sup>) | Pelvic_AntPost_Acc | Body kinematics |
| Peak thigh segment angular velocity in pre-swing (rad/s) | Thigh_Vel | Body kinematics |
| Peak shank segment angular velocity in pre-swing (rad/s) | Shank_Vel | Body kinematics |
| Relative peak shank angular velocity with respect to thigh (rad/s) | relShankThigh_Vel | Body kinematics |
| Paretic propulsive impulse in pre-swing<sup>6</sup> (%BW · s) | Propulsive_Impulse | Impulse |
| Paretic braking impulse in pre-swing<sup>6</sup> (%BW · s) | Braking_Impulse | Impulse |
| Paretic net impulse in pre-swing<sup>6</sup> (% BW · s) | Net_Impulse | Impulse |
| Peak anterior/posterior ground reaction force in pre-swing (% BW) | AntPost_GRF | GRF |
| Peak mediolateral GRF in stance (% BW) | MedLat_st_GRF | GRF |
| Peak vertical GRF in stance (% BW) | Vert_st_GRF | GRF |

\* BW: body weight

### Variable Selection

Variable selection is a procedure to select appropriate variables from a complete list of variables by removing those that are irrelevant or redundant. This step could be achieved by either parametric or nonparametric regression. The parametric approaches provide generalizable and interpretable statistical inference but require assumptions of linearity and normal distribution. Conversely, nonparametric machine learning methods do not need stringent distribution assumptions, but their models are less interpretable and risk overfitting without optimal tuning of hyper-parameters. In this study, we aimed to achieve a comprehensive variable selection by utilizing both parametric and nonparametric regression methods to address unknown variable relationships.

As a parametric approach, we deployed the least absolute shrinkage and selection operator (LASSO) method which is a penalized regression technique that uses an L1-norm penalty on the regression coefficients^40^. By the addition of the regularization term on linear regression, the LASSO regression produces a sparse and interpretable linear model with important variables automatically selected. In this study, we used *glmmLasso* package in R statistical software^16^ for modeling generalized linear mixed model using LASSO. We set a random intercept model with subjects as random effects and all predictors as fixed effects. We then built repeated 5-fold cross-validation to avoid a bias in the evaluation regression model. In data splitting, stratified sampling was applied based on subjects to obtain a sample population that best represents the entire population. In every cross-validation loop, the parameter λ was tuned by grid search using Bayesian Information Criteria (BIC). To quantify the importance of each predictor variable in regression, we ranked variables based on the selection proportions defined by frequencies of each selected variable in trained LASSO models from repeated cross-validation. The selection proportions have a score ranging from 0 (always shrunk, unimportant feature) to 1 (never shrunk, essential feature) by averaging variable selection frequencies so that we could infer individual feature importance in the regression model. The same ranking method was used in a previous regression study for post-stroke clinical outcomes^25^. In this study, we repeated 50 cross-validation repetitions to obtain variable selection proportions from the LASSO regression analysis. Variables above the threshold were determined as critical parameters for prediction. The threshold was 0.95 meaning 95% of the chance for selection across repeated cross-validation.

As a nonparametric approach, we deployed Bayesian Additive Regression Trees (BART), which is a nonparametric, ensemble method of regression tree models^9^. BART combines the advantages of the Bayesian model, incorporating past information about a parameter and forming a prior distribution for future analysis, and ensemble methods. It recently has gained popularity due to its superior prediction performance over other machine learning techniques (i.e., random forests, gradient boosting model, neural networks) in various study settings^9,19^. To evaluate the relative importance of each predictor variable from BART, Chipman et al. proposed the variable inclusion proportion corresponding to the proportion of times each variable is selected for splitting nodes across Markov chain Monte Carlo iterations in the sum-of-trees model^9^. Intuitively, variables with higher inclusion proportions are the more important variables in prediction. In this study, we implemented BART and extracted the variable inclusion proportions as a feature importance metric by using the *BART* package publicly available in R statistical software^36^. We performed a repeated 5-fold cross-validation with a total of 50 repetitions for unbiased regression outcomes. The stratified split dataset across repetitions was the same as the one used for glmmLASSO for a lateral comparison. Variables above the threshold, denoted by 1 / (total number of predictors), were determined as a crucial subset. This threshold stands for the probability of equal inclusion for all predictors in the model.

### Reduced Model Selection

With key variables resulting from both parametric and nonparametric variable selection, we fit reduced models using a linear mixed and BART regression. For linear mixed model regression, we formulated a random intercept model with nested random effects in terms of subjects and exoskeletal assistance. Parametric and nonparametric models were compared to each other based on goodness-of-fit measures, such as adjusted R^2^ and Root Mean Square Error (RMSE) acquired by 50 repetitions of 5-fold cross-validations. Based on the reduced fit model, we additionally performed a model selection through backward stepwise elimination. This stepwise regression eliminated insignificant variables resulting in the final best fit model with minimum numbers of predictors. Additionally, the relationships between variables were examined by Spearman’s rank correlation coefficient. The overall workflow was summarized in Figure 2.

**Figure 2:**
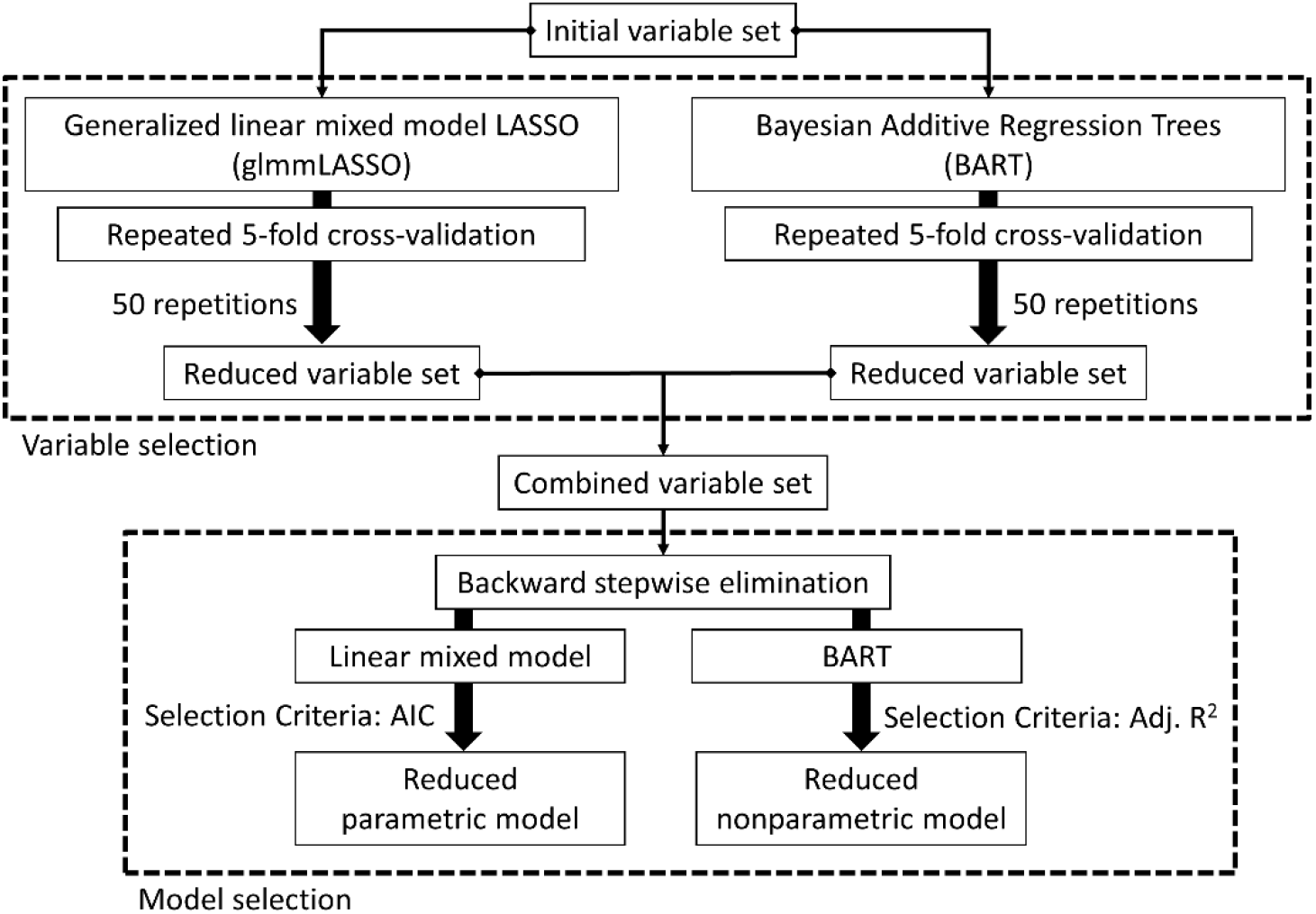
Overall workflow. AIC, Akaike information criterion; Adj. R^2^, Adjusted R^2^.

## Results

Parametric and non-parametric models showed similar results (**Figure 3**). Four parameters from kinematic and muscle-tendon state variables in the pre-swing phase remained (>95% of selection proportion) in the LASSO model across all 50 iterations: peak shank velocity, relative peak knee velocity with respect to hip, peak RF fiber stretch velocity, and peak knee flexion velocity. Using BART, six predictor variables from kinematic and muscle-tendon state variables in the pre-swing phase were determined as crucial variables. These were four common important variables between the parametric and nonparametric variable selection: the peak shank velocity, relative peak knee velocity with respect to the peak hip flexion, the peak RF fiber stretch velocity, and the peak knee flexion velocity. All parameters above the significance threshold using LASSO were also above the significance threshold using BART. Both variable selections resulted in low importance scores for all variables related to GRF and impulse.

**Figure 3:**
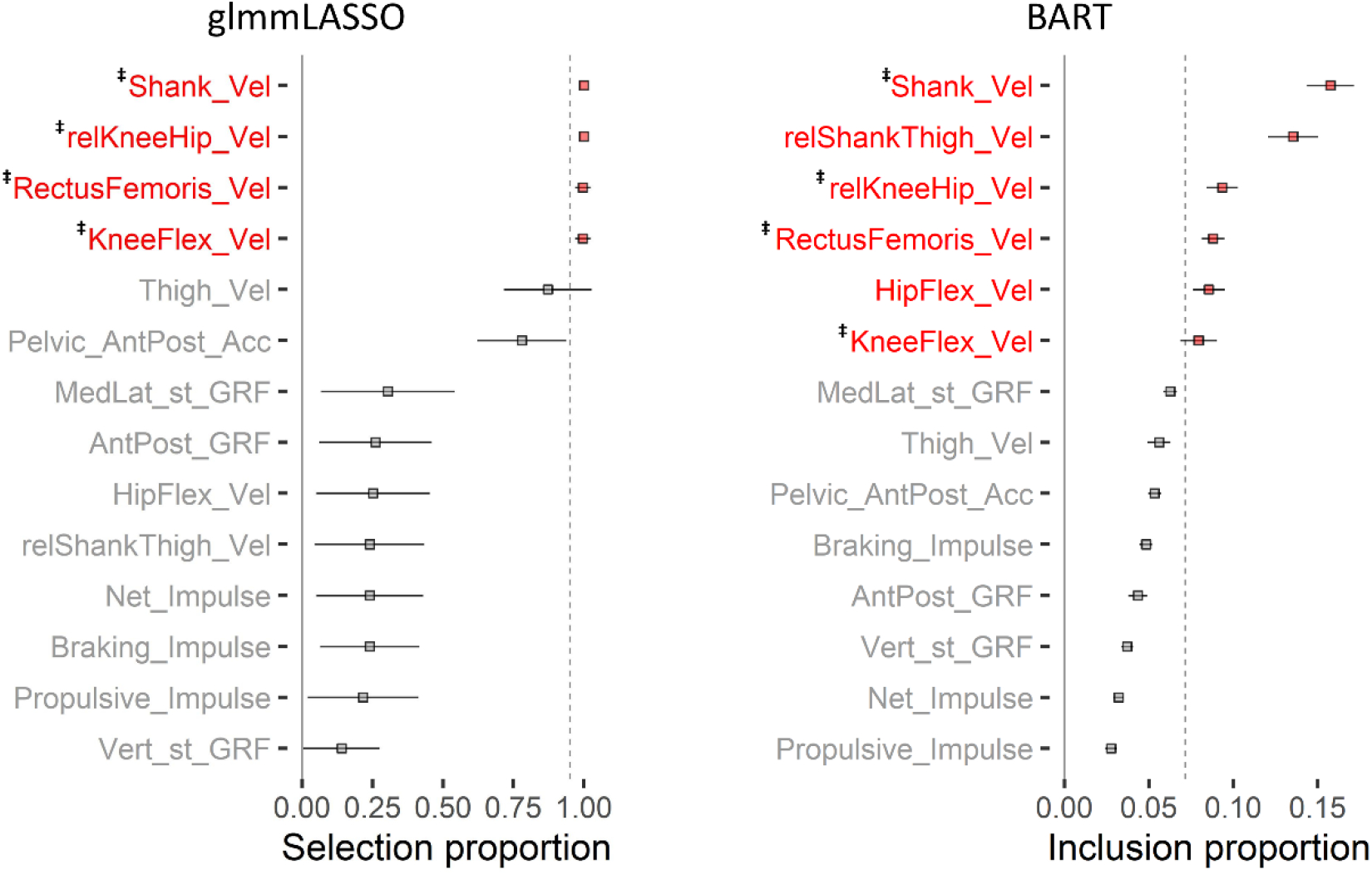
Variable selection results from glmmLASSO (left) and BART (right) for a total of 14 predictors. The square markers represent the average, and the solid lines are the standard deviation of total 50 repetitions. The red-colored variables are essential variables showing above threshold (vertical dashed line) denoted by 95% of selection proportions and 1/total number of variables for glmmLASSO and BART, respectively. The common essential variables between methods denoted by superscript symbol of ‡.

To ensure the model was both representative and not overly complex, we fitted a reduced model using all six significant predictors from **Figure 3**, and then evaluated the goodness-of-fit of models. We used the same random effects and the same hyperparameter setting as the variable selection. **Table 2** summarizes accuracy measures for full and reduced regression models from repeated 5-fold cross-validation. The reduced model showed slightly degraded goodness-of-fit (i.e., increased RMSE and decreased adjusted R^2^) compared to the full model. Except for the parametric regression model’s out-of-bag test, all accuracy measurements differed significantly between full and reduced models (*p* < .001).

**Table 2:**
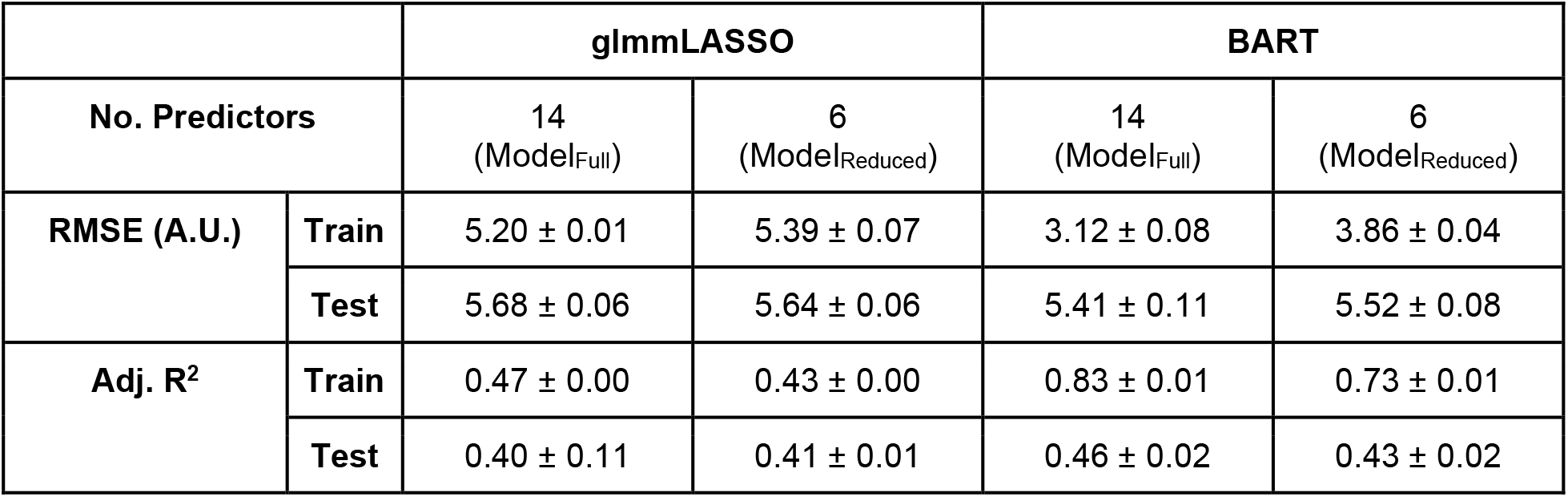
Summary of regression model accuracy (Mean ± SD) from repeated 5-fold cross-validation.

|  |  | glmLASSO |  | BART |  |
| --- | --- | --- | --- | --- | --- |
| No. Predictors |  | 14<br>(Model<sub>Full</sub>) | 6<br>(Model<sub>Reduced</sub>) | 14<br>(Model<sub>Full</sub>) | 6<br>(Model<sub>Reduced</sub>) |
| RMSE (A.U.) | Train | 5.20 ± 0.01 | 5.39 ± 0.07 | 3.12 ± 0.08 | 3.86 ± 0.04 |
| | Test | 5.68 ± 0.06 | 5.64 ± 0.06 | 5.41 ± 0.11 | 5.52 ± 0.08 |
| Adj. $R^2$ | Train | 0.47 ± 0.00 | 0.43 ± 0.00 | 0.83 ± 0.01 | 0.73 ± 0.01 |
| | Test | 0.40 ± 0.11 | 0.41 ± 0.01 | 0.46 ± 0.02 | 0.43 ± 0.02 |

Backward stepwise elimination additionally cut down the number of variables from the reduced model of six important predictors. The stepwise elimination on the parametric model was based on the Akaike information criterion (AIC). The adjusted R^2^ was used for the nonparametric model’s stepwise elimination. Stepwise regression using the parametric linear mixed model determined the last four crucial variables including two joint kinematic and another two body kinematic variables: peak hip flexion velocity, relative peak knee velocity to peak hip flexion, peak shank velocity and the relative peak shank velocity to peak thigh velocity. **Table 3** represents the summary of the final regression model by a random intercept mixed model with nested random effects with respect to subject and assistance. Small marginal R^2^ (0.074) but moderate conditional R^2^ (0.505) accounted for the most variance of this model. The adjusted R^2^ of this final model was 0.533. Backward elimination based on the nonparametric model, BART, revealed the same last four critical predictors. **Figure 4** illustrates the goodness-of-fit changes through backward elimination using BART. Once the model consisted of less than four predictors, its adjusted R^2^ started decreasing significantly meaning that variables in the gray shaded area of **Figure 4** were the minimum crucial predictor set. The adjusted R^2^ of BART model with four variables was 0.638. The results were robust to differences in timing (**Supplementary material**).

**Table 3:**
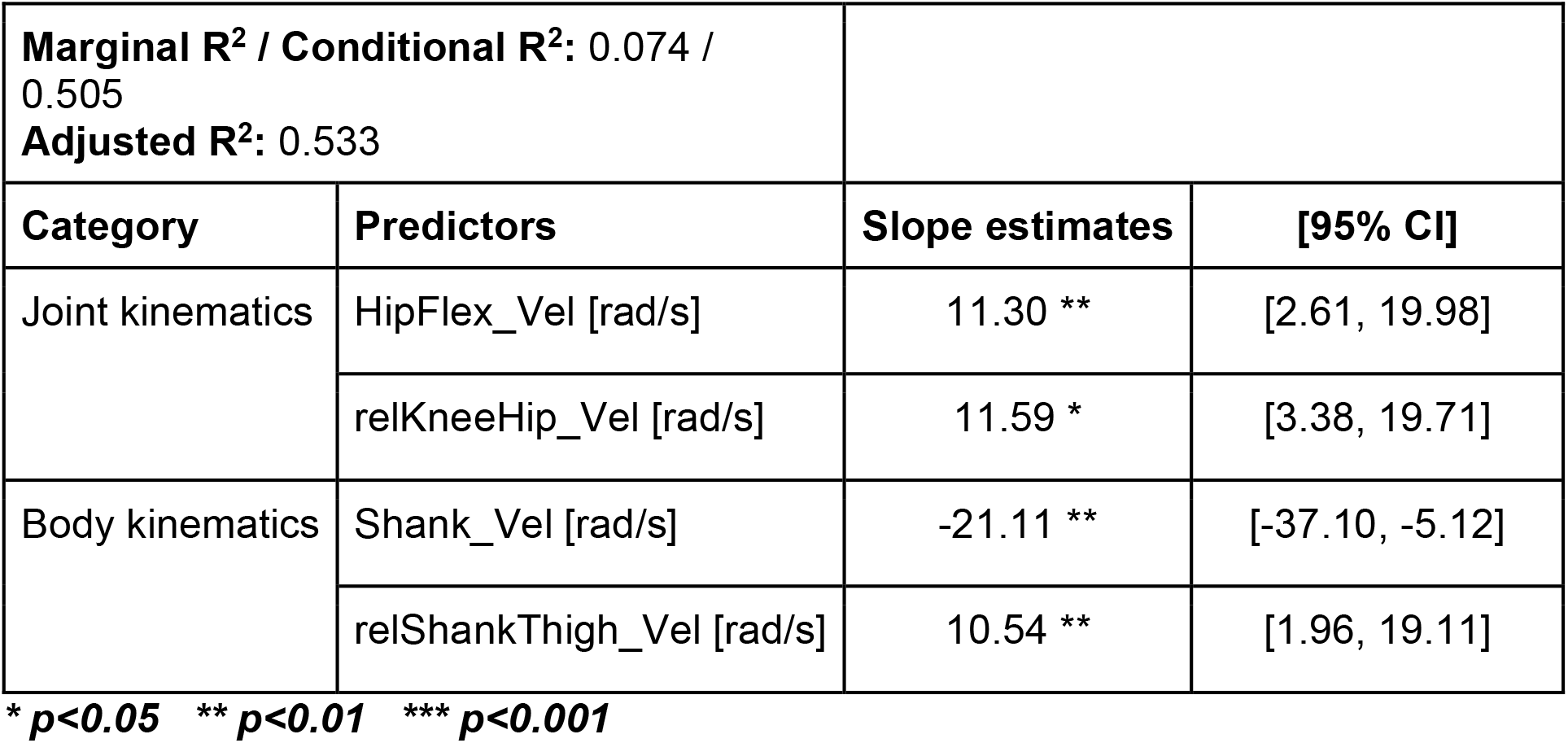
Summary of final model from stepwise backward regression.

| **Marginal $R^2$ / Conditional $R^2$: 0.074 / 0.505** |  |  |  |
| --- | --- | --- | --- |
| **Adjusted $R^2$: 0.533** |  |  |  |
| **Category** | **Predictors** | **Slope estimates** | **[95% CI]** |
| Joint kinematics | HipFlex_Vel [rad/s] | 11.30 ** | [2.61, 19.98] |
|  | relKneeHip_Vel [rad/s] | 11.59 * | [3.38, 19.71] |
| Body kinematics | Shank_Vel [rad/s] | -21.11 ** | [-37.10, -5.12] |
|  | relShankThigh_Vel [rad/s] | 10.54 ** | [1.96, 19.11] |

\* $p < 0.05$ \*\* $p < 0.01$ \*\*\* $p < 0.001$

**Figure 4:**
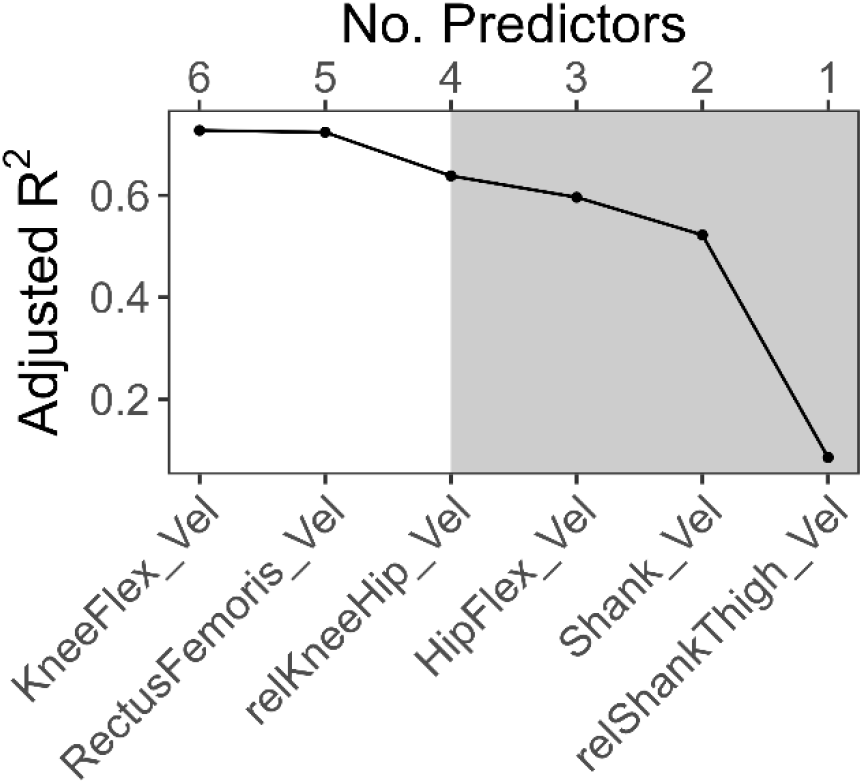
Goodness-of-fit changes by BART stepwise backward elimination. The variable names shown in the bottom axis are the predictors removed at each step. For example, “KneeFlex_Vel” was removed at the first step of elimination from a model with six variables. The gray shaded area indicates the steps that the adjusted R^2^ was significantly affected by the elimination.

The relationships between selected kinematic predictors and muscle-tendon state, pre-swing rectus femoris fiber stretch velocity, were examined by Spearman’s rank correlation coefficient. **Table 4** summarizes the coefficients.

**Table 4:** Spearman’s rank correlation coefficient (ρ) between key kinematic predictors and pre-swing rectus femoris fiber stretch velocity.

| Category | Predictors | $\rho$ |
| --- | --- | --- |
| Joint kinematics | HipFlex_Vel [rad/s] | 0.59 *** |
|  | relKneeHip_Vel [rad/s] | 0.90 *** |
| Body kinematics | Shank_Vel [rad/s] | 0.87 *** |
|  | relShankThigh_Vel [rad/s] | -0.18 *** |

\* $p < 0.05$ \*\* $p < 0.01$ \*\*\* $p < 0.001$

## Discussion

Exoskeletal assistance has the potential to unload some of the effort from clinicians and caregivers but does not yet account for other neuromuscular impairments such as hyperreflexia^1,37^. The aim of this study was to determine accessible biomechanical predictors of quadriceps hyperreflexia based on walking data from those with post-stroke SKG with and without knee flexion exoskeletal assistance. We used both parametric and non-parametric regression techniques to obtain two different perspectives of the key kinematic, kinetic and muscle-tendon predictors of rectus femoris hyperreflexia. We found four kinematic predictors obtained during pre-swing phase were shared between the regression techniques: peak hip flexion velocity, the relative peak knee to peak hip flexion velocity, peak shank angular velocity, and the relative peak shank to peak thigh angular velocity. These parameters can be measured by two or three wearable sensors. The implication of these findings is that these parameters can be used to predict hyperreflexia and control during exoskeletal assistance.

Spasticity is often characterized as velocity-dependent hyperexcitability of the stretch reflex^23^. Since it was widely believed that the spindle proprioceptive receptors encode information for muscle length and velocity changes^20,26^, spasticity modeling has been based on muscle length and velocity feedback^24^. Our group’s previous musculoskeletal simulation analysis confirmed the strong association between simulated RF muscle fiber stretch velocity and RF excitability^2^. In this study, both parametric and nonparametric regression techniques for variable selection resulted in the same finding that kinematic variables are predictive of an RF reflex. Indeed, there were strong correlations (ρ ≃ 0.90) between some of the selected predictors (relative peak knee flexion velocity to peak hip flexion velocity, peak shank angular velocity) and RF fiber stretch velocity (**Table 4**), as we expected. We did not expect that these parameters would be more predictive of a reflex than RF fiber stretch velocity, the physiological mechanism of a stretch reflex. The fiber stretch velocity was computed using generic Hill-type muscle models and mathematically driven cost functions^39^, rather than direct measurement, a model validated in our previous work^2^. Yet, it remains possible that RF fiber stretch velocity could have played a stronger statistical role if the true value could have been obtained. Regardless of the level of significance between variables, it is notable that out of all 14 parameters, the ones found to be most predictive of a reflex were ones related to hip and knee kinematics as well as RF fiber stretch velocity. This finding is consistent with our hypothesis that more easily measurable kinematic variables can serve as a proxy for fiber stretch velocity in exoskeletal control.

Over the past decade, the exoskeleton community has moved towards human-in-the-loop design^43^, including the incorporation of real-time estimation of musculotendon mechanics^32^, and ultrasound imaging^13,29,35^. While these approaches provide an exciting potential for the future of personalized robotics, the extra equipment remains limited to a few research institutes and adds significant complexity. In this work we found that joint kinematics accurately predict reflex responses in the RF. We estimate that kinematic predictors from this study could be measured by only two or three portable motion sensors such as inertial measurement units. This is significant because it questions the basis for using complex imaging or computational technology in lieu of simpler proxies. Enabling the use of ubiquitous sensor systems would allow exoskeleton engineers and rehabilitation researchers to design practical and effective devices.

It is possible that this work could lack a wide range of variables. Due to the exponential increase in the sampling volume, adding extra dimensionality is not always beneficial in regression analysis, specifically when there are sparse relevant predictors compared to the total number of predictors or the fundamental relationships are nonlinear (so called “curse of dimensionality”)^14^. To avoid these adverse effects, we limited the scope of parameter sources among previously assessed associations with post-stroke SKG. Additionally, we did not include parameters that were unable to be directly measured, such as joint kinetics, except for simulated muscle fiber stretch velocity. This omission was due to the practical difficulty in measuring real time joint kinetics. Therefore, in this work, we aimed to balance comprehensiveness, practicality and overfitting.

This study has several limitations. Regression approaches based on observations from only 8 participants may affect generalization of the results. Despite the small sample size, the repeated 5-fold cross-validation^34^ took into account the randomness in sampling resulting in unbiased and trustworthy variable and model selection results. This study does not provide causal relationships between kinematic parameters and quadriceps hyperreflexia. However, it is notable that the predictors of RF hyperreflexia (i.e. related to knee-hip velocities) found in this work were all physiologically related to the biarticular span of the RF suggesting that this statistical approach may help reveal mechanisms of hyperreflexia. For instance, the relative peak angular velocity between the knee and hip was the most correlated variable with RF fiber stretch velocity (ρ = 0.90 in **Table 4**). Still, the relative peak angular velocity between the shank and thigh had a weak negative correlation (ρ = -0.18 in **Table 4**) to RF fiber stretch velocity. While thigh-shank segment and knee-hip joint velocities are not equivalent, the large difference in correlations is difficult to explain mechanistically and other approaches may be needed to investigate further. Lastly, our findings were based on two types of regression techniques among numerous statistical and machine learning approaches. Although there exists an advanced penalized regression approach improving the limitations of LASSO^33^ (i.e., Elastic Net^44^), glmmLASSO in this study could result in a better model due to subjects’ random-effects modeling. There were also popular tree-based machine learning algorithms, such as gradient boosting^28^ and random forest methods^5^. It has been reported that BART has outperformed these other methods^9^. Therefore, the use of these two different regression methods was sufficient.

## Conclusions

Despite the rapid growth of wearable robot technology, there is still a lack of consideration regarding neuromuscular impairments such as hyperreflexia into device design. In this study, we introduced regression-based variable selection for predicting rectus femoris hyperreflexia following pre-swing knee flexion exoskeletal assistance. We found that four kinematic variables relevant to knee and hip joint motions were sufficient to effectively predict hyperreflexia in people with post-stroke SKG. These results suggest that effective monitoring and control of these accessible knee and hip kinematics may be more productive at regulating hyperreflexia than the more complex acquisition of muscle fiber stretch velocity. This work represents a novel statistical approach to identifying biomechanical associations with reflex function. This information could be used to further understand the mechanisms of hyperreflexia and its associated clinical interventions.

## Data Availability

All data produced in the present study are available upon reasonable request to the authors

## Funding

This work was supported by the National Center for Medical Rehabilitation Research at the National Institute of Child Health and Human Development under the award number R01HD100416.

## Supplementary Material

We performed a sensitivity analysis based on two additional reflex response windows by changing time windows for computing involuntary RF muscle activity: 90 ms and 150 ms. Figures A1, A2 and Table A1 summarize variable selection for 90 ms time window for a reflex response and Figures A3, A4 and Table A2 are for 150 ms. The last four key selected variables were invariant regarding the timing error of RF fiber stretch velocity estimated by OpenSim simulation.

**Figure A1:**
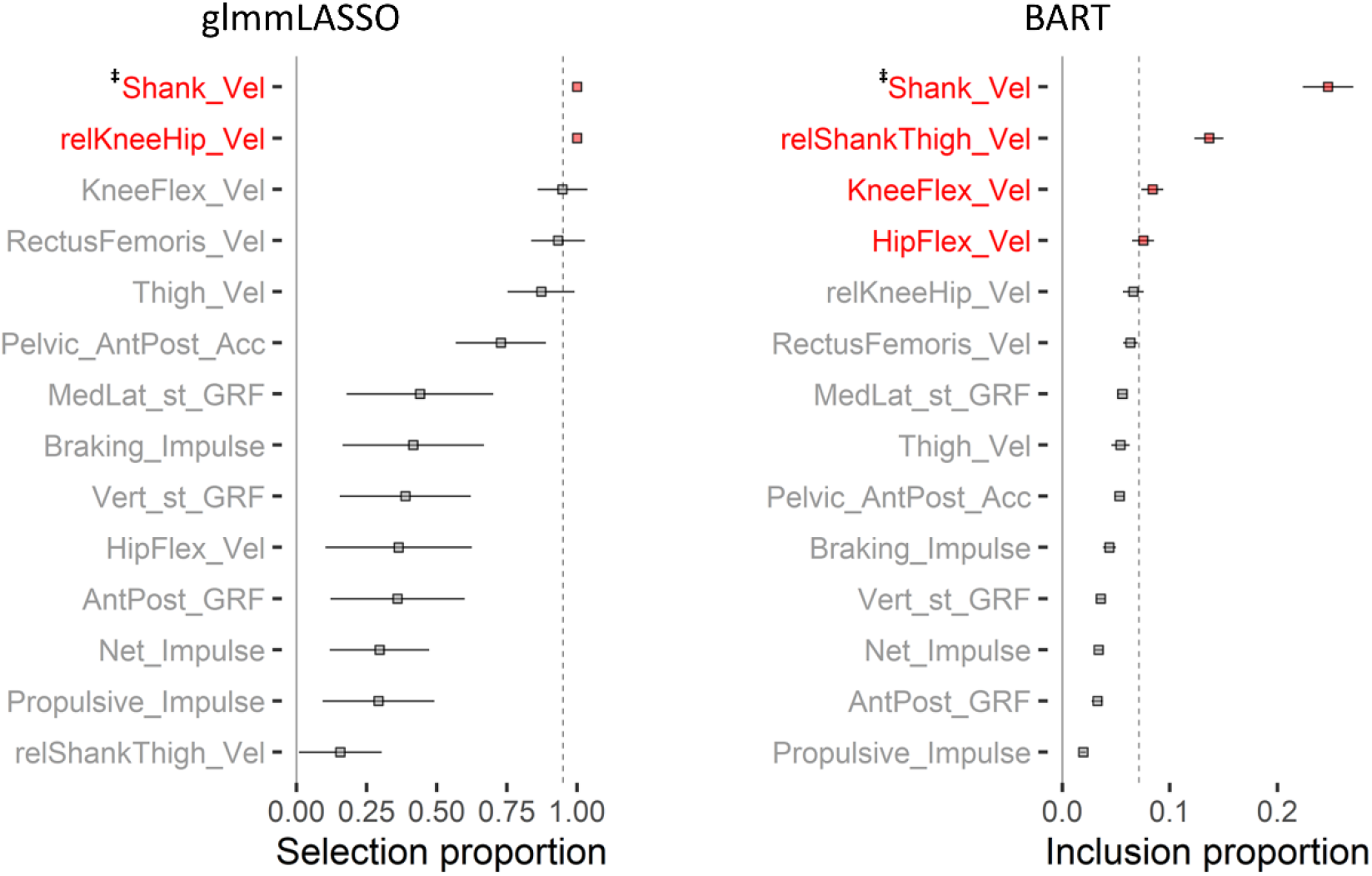
Variable selection results from 90 ms time-window of a reflex response. The left figure was from glmmLASSO and the right figure from BART.

**Figure A2:**
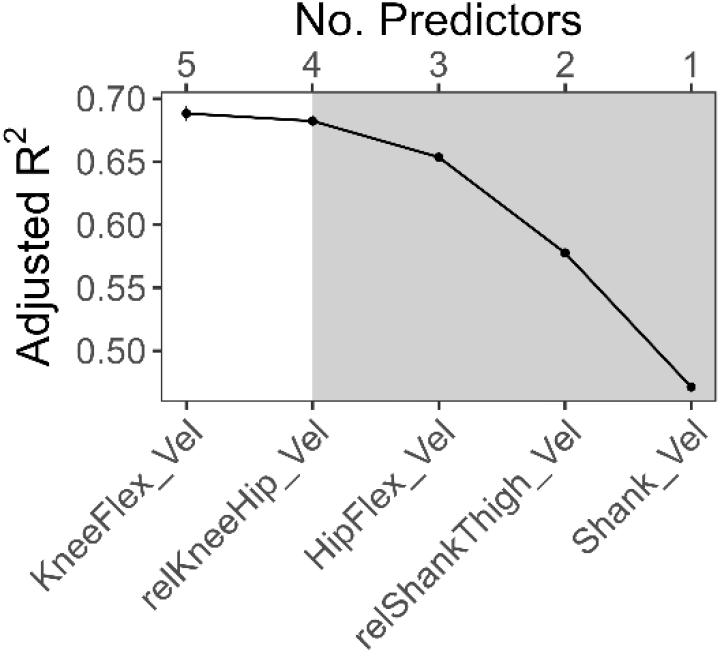
Goodness-of-fit changes by BART stepwise backward elimination for 90 ms time-window of a reflex response.

**Table A1:**
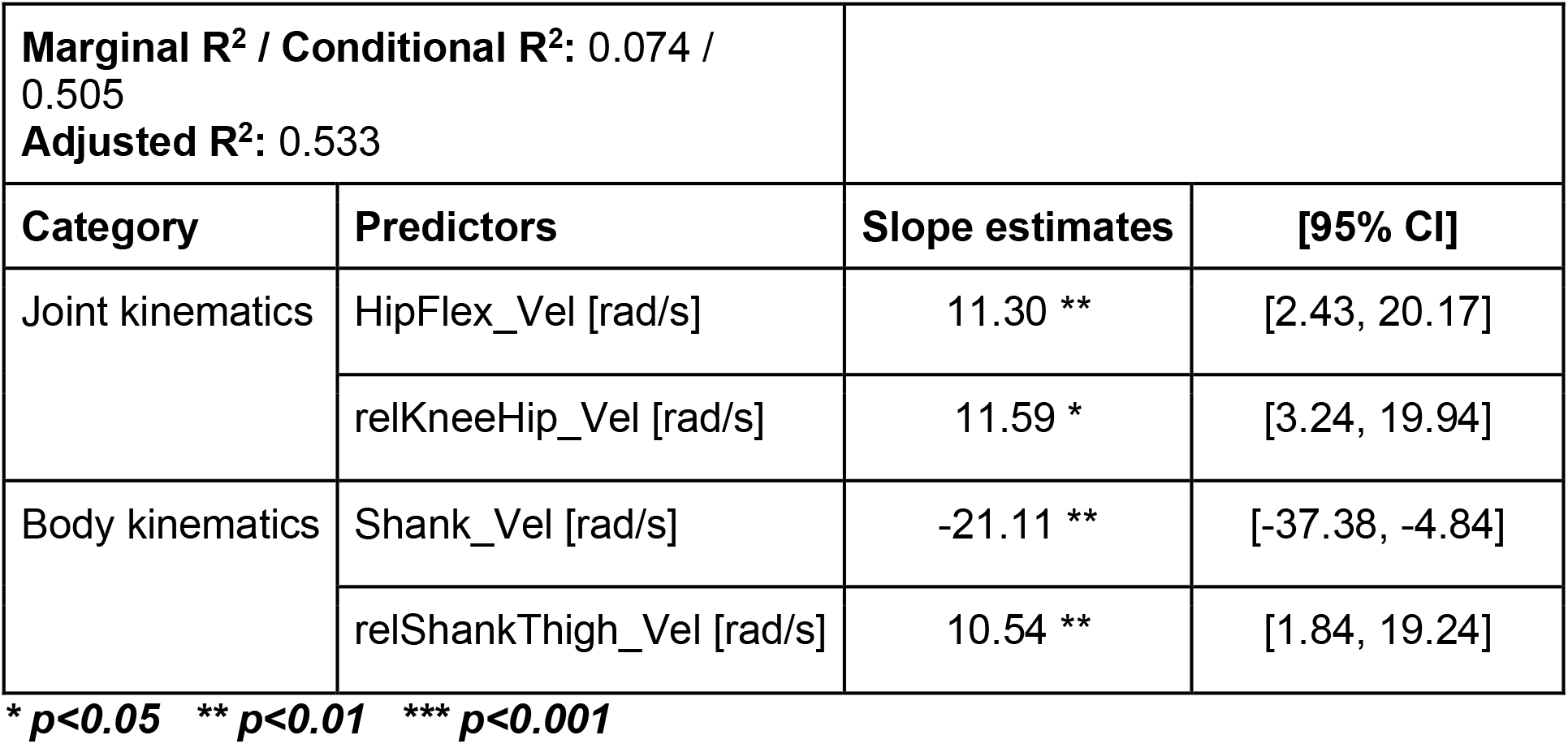
Summary of final model from stepwise backward regression for 90 ms time-window of a reflex response.

| **Marginal R<sup>2</sup> / Conditional R<sup>2</sup>: 0.074 / 0.505**<br>**Adjusted R<sup>2</sup>: 0.533** |  |  |  |
| --- | --- | --- | --- |
| Category | Predictors | Slope estimates | [95% CI] |
| Joint kinematics | HipFlex_Vel [rad/s] | 11.30 ** | [2.43, 20.17] |
|  | relKneeHip_Vel [rad/s] | 11.59 * | [3.24, 19.94] |
| Body kinematics | Shank_Vel [rad/s] | -21.11 ** | [-37.38, -4.84] |
|  | relShankThigh_Vel [rad/s] | 10.54 ** | [1.84, 19.24] |

\* $p < 0.05$ \*\* $p < 0.01$ \*\*\* $p < 0.001$

**Figure A3:**
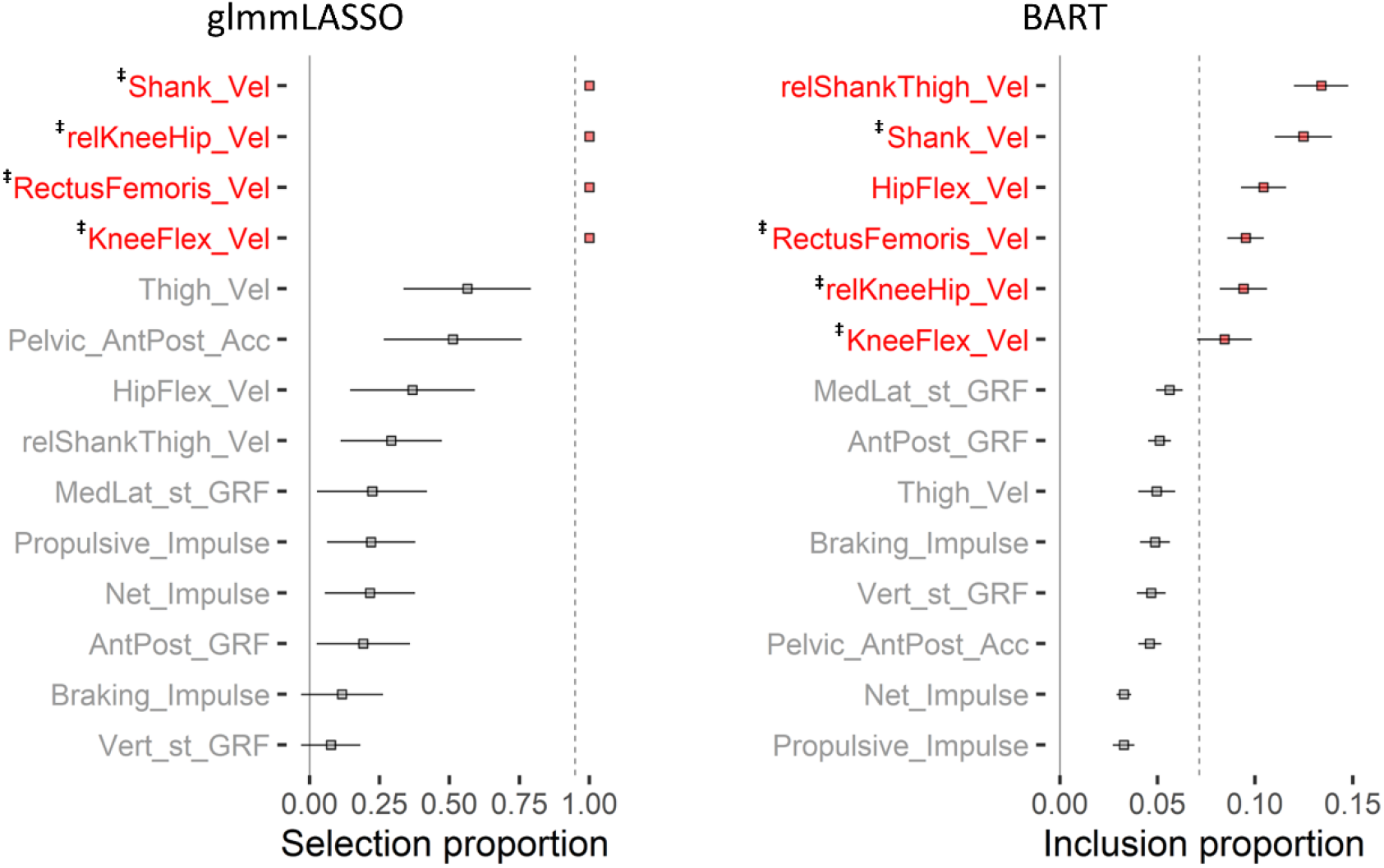
Variable selection results from 150 ms time-window of a reflex response. The left figure was from glmmLASSO and the right figure from BART.

**Figure A4:**
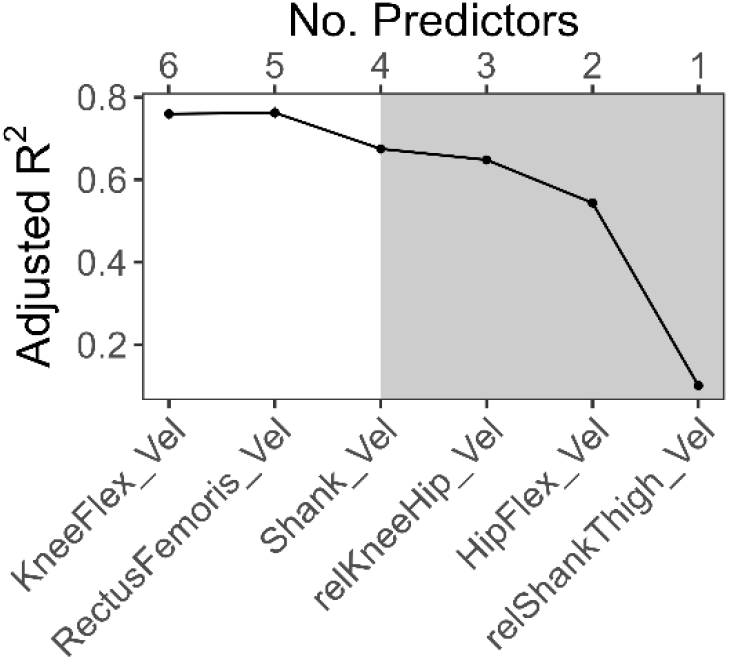
Goodness-of-fit changes by BART stepwise backward elimination for 150 ms time-window of a reflex response.

**Table A2:**
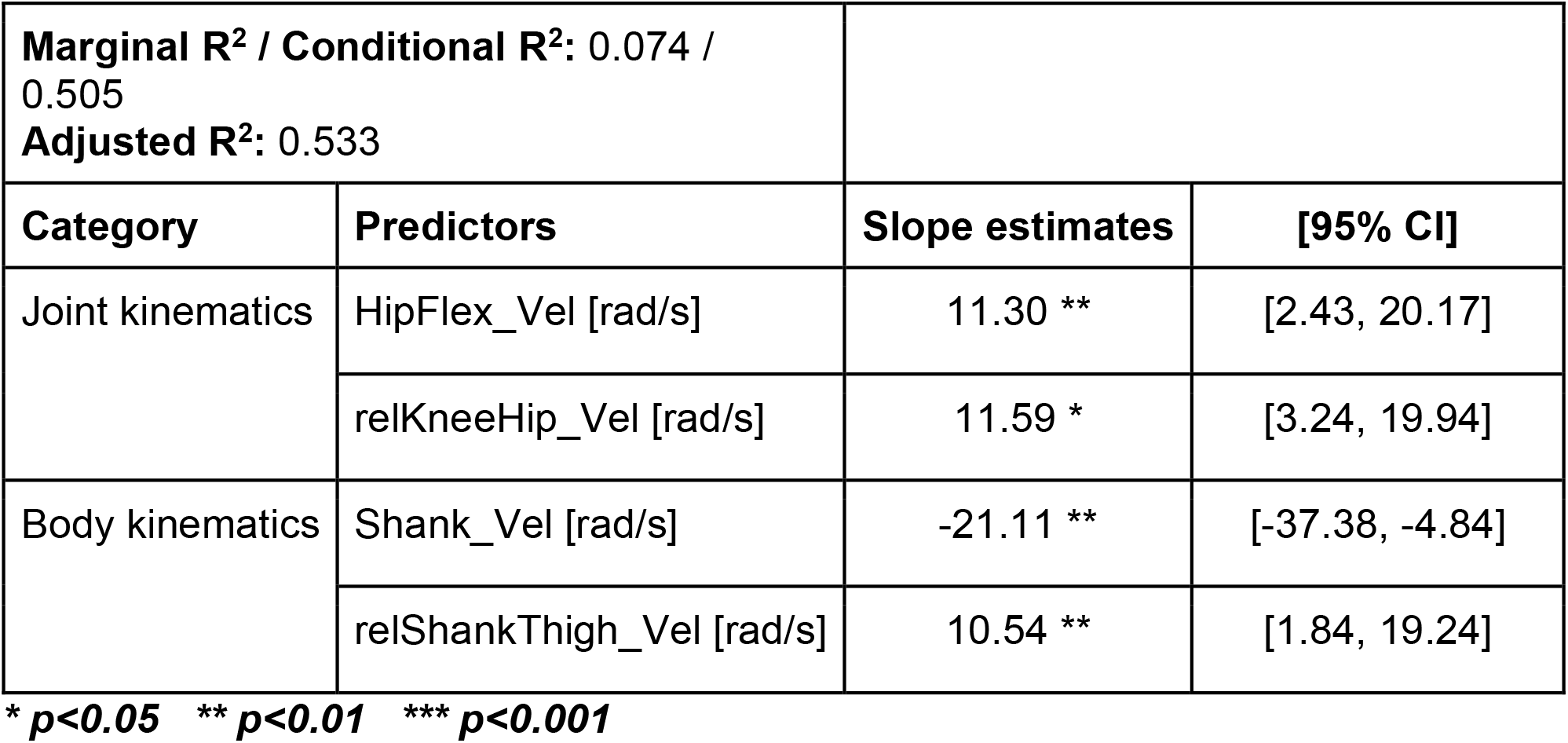
Summary of final model from stepwise backward regression for 150 ms time-window of a reflex response.

## Notes

### Competing Interest Statement

The authors have declared no competing interest.

### Author Declarations

University of Texas at Austin has waived ethics approval because it used anonymized data previously collected

## References

1. Akbas, T., K. Kim, K. Doyle, K. Manella, R. Lee, P. Spicer, M. Knikou, and J. Sulzer. Rectus femoris hyperreflexia contributes to Stiff-Knee gait after stroke. Journal of NeuroEngineering and Rehabilitation 17:117, 2020.

2. Akbas, T., R. R. Neptune, and J. Sulzer. Neuromusculoskeletal Simulation Reveals Abnormal Rectus Femoris-Gluteus Medius Coupling in Post-stroke Gait. Frontiers in Neurology 10:301, 2019.

3. Akbas, T., and J. Sulzer. Implementing a virtual gait assistance device within a musculoskeletal simulation framework., 2015.

4. Akbas, T., and J. Sulzer. Musculoskeletal simulation framework for impairment-based exoskeletal assistance post-stroke., 2019.doi:10.1109/ICORR.2019.8779564

5. Biau, G., and E. Scornet. A random forest guided tour. TEST 25:197–227, 2016.

6. Bowden, M. G., C. K. Balasubramanian, R. R. Neptune, and S. A. Kautz. Anterior-Posterior Ground Reaction Forces as a Measure of Paretic Leg Contribution in Hemiparetic Walking. Stroke 37:872–876, 2006.

7. Bowden, M. G., A. L. Behrman, M. Woodbury, C. M. Gregory, C. A. Velozo, and S. A. Kautz. Advancing Measurement of Locomotor Rehabilitation Outcomes to Optimize Interventions and Differentiate between Recovery versus Compensation. Journal of neurologic physical therapy: JNPT 36:38, 2012.

8. Burden, A. How should we normalize electromyograms obtained from healthy participants? What we have learned from over 25years of research. Journal of Electromyography and Kinesiology 20:1023–1035, 2010.

9. Chipman, H. A., E. I. George, and R. E. McCulloch. BART: Bayesian additive regression trees. The Annals of Applied Statistics 4:266–298, 2010.

10. Delp, S. L., F. C. Anderson, A. S. Arnold, P. Loan, A. Habib, C. T. John, E. Guendelman, and D. G. Thelen. OpenSim: Open-Source Software to Create and Analyze Dynamic Simulations of Movement. IEEE Transactions on Biomedical Engineering 54:1940–1950, 2007.

11. Duncan, P. W., R. Zorowitz, B. Bates, J. Y. Choi, J. J. Glasberg, G. D. Graham, R. C. Katz, K. Lamberty, and D. Reker. Management of Adult Stroke Rehabilitation Care. Stroke 36:e100–e143, 2005.

12. Esquenazi, A., M. Talaty, and A. Jayaraman. Powered Exoskeletons for Walking Assistance in Persons with Central Nervous System Injuries: A Narrative Review. PM&R 9:46–62, 2017.

13. Franz, J. R. A sound approach to improving exoskeletons and exosuits. Science Robotics, 2021.doi:10.1126/scirobotics.abm6369

14. Geenens, G. Curse of dimensionality and related issues in nonparametric functional regression. Statistics Surveys 5:30–43, 2011.

15. Goldberg, S. R., S. Õunpuu, A. S. Arnold, J. R. Gage, and S. L. Delp. Kinematic and kinetic factors that correlate with improved knee flexion following treatment for stiff-knee gait. Journal of Biomechanics 39:689–698, 2006.

16. Groll, A., and G. Tutz. Variable selection for generalized linear mixed models by L 1-penalized estimation. Stat Comput 24:137–154, 2014.

17. Hendricks, H. T., J. van Limbeek, A. C. Geurts, and M. J. Zwarts. Motor recovery after stroke: A systematic review of the literature. Archives of Physical Medicine and Rehabilitation 83:1629–1637, 2002.

18. Hicks, J. L., T. K. Uchida, A. Seth, A. Rajagopal, and S. L. Delp. Is My Model Good Enough? Best Practices for Verification and Validation of Musculoskeletal Models and Simulations of Movement. Journal of Biomechanical Engineering 137:, 2015.

19. Hill, J. L. Bayesian Nonparametric Modeling for Causal Inference. Journal of Computational and Graphical Statistics 20:217–240, 2011.

20. Houk, J. C., W. Z. Rymer, and P. E. Crago. Dependence of dynamic response of spindle receptors on muscle length and velocity. Journal of Neurophysiology 46:143–166, 1981.

21. Jørgensen, H. S., H. Nakayama, H. O. Raaschou, and T. S. Olsen. Recovery of walking function in stroke patients: The copenhagen stroke study. Archives of Physical Medicine and Rehabilitation 76:27–32, 1995.

22. Kalita, B., J. Narayan, and S. K. Dwivedy. Development of Active Lower Limb Robotic-Based Orthosis and Exoskeleton Devices: A Systematic Review. Int J of Soc Robotics 13:775–793, 2021.

23. Lance, J. W. The control of muscle tone, reflexes, and movement: Robert Wartenbeg Lecture. Neurology 30:1303–1303, 1980.

24. Le Cavorzin, P., S. A. Poudens, F. Chagneau, G. Carrault, H. Allain, and P. Rochcongar. A comprehensive model of spastic hypertonia derived from the pendulum test of the leg. Muscle & Nerve 24:1612–1621, 2001.

25. Majeed, Y. A., S. S. Awadalla, and J. L. Patton. Regression techniques employing feature selection to predict clinical outcomes in stroke. PLOS ONE 13:e0205639, 2018.

26. Matthews, P. B. C. The response of de-efferented muscle spindle receptors to stretching at different velocities. J Physiol 168:660–678, 1963.

27. Mrachacz-Kersting, N., B. A. Lavoie, J. B. Andersen, and T. Sinkjaer. Characterisation of the quadriceps stretch reflex during the transition from swing to stance phase of human walking. Exp Brain Res 159:108–122, 2004.

28. Natekin, A., and A. Knoll. Gradient boosting machines, a tutorial. Frontiers in Neurorobotics 7:, 2013.

29. Nuckols, R. W., S. Lee, K. Swaminathan, D. Orzel, R. D. Howe, and C. J. Walsh. Individualization of exosuit assistance based on measured muscle dynamics during versatile walking. Science Robotics, 2021.doi:10.1126/scirobotics.abj1362

30. Piazza, S. J., and S. L. Delp. The influence of muscles on knee flexion during the swing phase of gait. Journal of Biomechanics 29:723–733, 1996.

31. Pierrot-Deseilligny, E., and D. Burke. The circuitry of the human spinal cord: its role in motor control and movement disorders. Cambridge university press, 2005.

32. Pizzolato, C., D. G. Lloyd, M. Sartori, E. Ceseracciu, T. F. Besier, B. J. Fregly, and M. Reggiani. CEINMS: A toolbox to investigate the influence of different neural control solutions on the prediction of muscle excitation and joint moments during dynamic motor tasks. Journal of Biomechanics 48:3929–3936, 2015.

33. Ranstam, J., and J. A. Cook. LASSO regression. British Journal of Surgery 105:1348, 2018.

34. de Rooij, M., and W. Weeda. Cross-Validation: A Method Every Psychologist Should Know. Advances in Methods and Practices in Psychological Science 3:248–263, 2020.

35. Sheng, Z., A. Iyer, Z. Sun, K. Kim, and N. Sharma. A Hybrid Knee Exoskeleton Using Real-Time Ultrasound-Based Muscle Fatigue Assessment. IEEE/ASME Transactions on Mechatronics 1–9, 2022.doi:10.1109/TMECH.2022.3171086

36. Sparapani, R., C. Spanbauer, and R. McCulloch. Nonparametric Machine Learning and Efficient Computation with Bayesian Additive Regression Trees: The BART R Package. Journal of Statistical Software 97:1–66, 2021.

37. Sulzer, J. S., K. E. Gordon, Y. Y. Dhaher, M. A. Peshkin, and J. L. Patton. Preswing Knee Flexion Assistance Is Coupled With Hip Abduction in People With Stiff-Knee Gait After Stroke. Stroke 41:1709–1714, 2010.

38. Sulzer, J. S., R. A. Roiz, M. A. Peshkin, and J. L. Patton. A Highly Backdrivable, Lightweight Knee Actuator for Investigating Gait in Stroke. IEEE Transactions on Robotics 25:539–548, 2009.

39. Thelen, D. G., and F. C. Anderson. Using computed muscle control to generate forward dynamic simulations of human walking from experimental data. Journal of Biomechanics 39:1107–1115, 2006.

40. Tibshirani, R. Regression Shrinkage and Selection Via the Lasso. Journal of the Royal Statistical Society: Series B (Methodological) 58:267–288, 1996.

41. Young, A. J., and D. P. Ferris. State of the Art and Future Directions for Lower Limb Robotic Exoskeletons. IEEE Transactions on Neural Systems and Rehabilitation Engineering 25:171–182, 2017.

42. Young, R. R. Spasticity: a review. Neurology 44:S12–20, 1994.

43. Zhang, J., P. Fiers, K. A. Witte, R. W. Jackson, K. L. Poggensee, C. G. Atkeson, and S. H. Collins. Human-in-the-loop optimization of exoskeleton assistance during walking. Science, 2017.doi:10.1126/science.aal5054

44. Zou, H., and T. Hastie. Regularization and variable selection via the elastic net. Journal of the Royal Statistical Society: Series B (Statistical Methodology) 67:301–320, 2005.

